# Neuropathic symptoms with SARS-CoV-2 vaccination

**DOI:** 10.1101/2022.05.16.22274439

**Authors:** Farinaz Safavi, Lindsey Gustafson, Brian Walitt, Tanya Lehky, Sara Dehbashi, Amanda Wiebold, Yair Mina, Susan Shin, Baohan Pan, Michael Polydefkis, Anne Louise Oaklander, Avindra Nath

## Abstract

**Background and Objectives:** Various peripheral neuropathies, particularly those with sensory and autonomic dysfunction may occur during or shortly after acute COVID-19 illnesses. These appear most likely to reflect immune dysregulation. If similar manifestations can occur with the vaccination remains unknown.

**Results:** In an observational study, we studied 23 patients (92% female; median age 40years) reporting new neuropathic symptoms beginning within 1 month after SARS-CoV-2 vaccination. 100% reported sensory symptoms comprising severe face and/or limb paresthesias, and 61% had orthostasis, heat intolerance and palpitations. Autonomic testing in 12 identified seven with reduced distal sweat production and six with positional orthostatic tachycardia syndrome. Among 16 with lower-leg skin biopsies, 31% had diagnostic/subthreshold epidermal neurite densities (≤5%), 13% were borderline (5.01-10%) and 19% showed abnormal axonal swelling. Biopsies from randomly selected five patients that were evaluated for immune complexes showed deposition of complement C4d in endothelial cells. Electrodiagnostic test results were normal in 94% (16/17). Together, 52% (12/23) of patients had objective evidence of small-fiber peripheral neuropathy. 58% patients (7/12) treated with oral corticosteroids had complete or near-complete improvement after two weeks as compared to 9% (1/11) of patients who did not receive immunotherapy having full recovery at 12 weeks. At 5-9 months post-symptom onset, 3 non-recovering patients received intravenous immunoglobulin with symptom resolution within two weeks.

**Conclusions:** This observational study suggests that a variety of neuropathic symptoms may manifest after SARS-CoV-2 vaccinations and in some patients might be an immune-mediated process.

## Introduction

Vaccination against SARS-CoV-2 is the most important public health strategy to control the COVID-19 pandemic and decrease mortality and morbidity from the infection. The safety profile of current FDA approved vaccines characterizes a small number of post-immunization side effects. Vaccine roll-out was initiated in the United States in Dec 2020 and reports of adverse events to the Vaccine Adverse Event Reporting System include a wide variety of neurological and systemic manifestations.^1^

Such events are identified following large-scale vaccination campaigns and they may share immunological mechanisms with post-infectious neurological complications. Importantly, immune-mediated neurological adverse events post-vaccination are rare and often less severe than those that follow actual infection. A series of patients with a rare variant of Guillain-Barré syndrome with facial diplegia and paresthesia following the first dose of the AstraZeneca vaccine have been reported,^2^ and 3 of 11,636 participants in the ChAdOx1 nCoV-19 vaccine (AZD1222) trial developed transverse myelitis.^3^ One case of new onset small fiber neuropathy (SFN) and a second of postural orthostatic tachycardia syndrome (POTS) in previously healthy individuals were reported after receiving mRNA vaccines to SARS-CoV-2.^4-6^

Autoimmune autonomic neuropathy and ganglionopathy is an antibody-mediated disease that usually presents with orthostasis and symptoms of cholinergic failure including dry mouth or urinary retention.^7^ Sensory and autonomic neuropathies also occur following viral infections and vaccination without cholinergic manifestations or characterized autoantibodies. Regardless of whether defined autoantibodies were detected, symptoms and biomarkers often respond to high dose corticosteroid treatment suggesting the process is immune-mediated.^7^ Since onset of the COVID-19 pandemic, orthostatic intolerance, POTS, and peripheral neuropathies, have been observed as post-acute neurological sequelae of COVID-19.^8, 9^ Further investigation is required to explore underlying mechanisms and targeted therapies for these neurologic disorders.

Here, we report clinical evaluations of patients with new onset paresthesias with or without autonomic symptoms incident to SARS-CoV-2 vaccination and response to immunotherapy with corticosteroids or intravenous immunoglobulin (IVIg).

## Methods

Twenty-three self-referred patients were evaluated between January to September 2021 for new onset of potential symptoms of polyneuropathy (sensory, motor, or autonomic) within 1 month of SARS-CoV-2 vaccination were enrolled after consent to an IRB approved study at the National Institutes of Health (protocol # 15-N-00125). All were evaluated by in-person (n=13) and/or televisit (n=10), and data was abstracted from their medical records. We excluded patients whose complaints were non-neurologic or limited to worsening or recrudescence of prior neurological symptoms and those with underlying conditions or risk factors for neuropathy and dysautonomia. All patients were extensively investigated for other causes of SFN. Two had organ-specific autoimmune disorders (one with Crohn’s disease, one with Hashimoto thyroiditis) in clinical remission at the time of vaccination. Table 1 reports participants’ demographic characteristics.

**Table 1:** Demographic characteristics

| Demographic Information | Case# 1* | 2 | 3* | 4 | 5 | 6* | 7 | 8* | 9* | 10 | 11* |
| --- | --- | --- | --- | --- | --- | --- | --- | --- | --- | --- | --- |
| Age Range | 31-40 | 61-70 | 31-40 | 31-40 | 51-60 | 61-70 | 61-70 | 51-60 | 51-60 | 31-40 | 31-40 |
| Sex | F | F | F | F | F | F | F | F | M | F | F |
| Type of vaccine | AstraZeneca | Pfizer | Pfizer | Pfizer | Moderna | Pfizer | Pfizer | Moderna | Janssen | Moderna | Moderna |
| Dose | 1st | 1<sup>st</sup> | 2nd | 1st | 1st | 2nd | 1st | 1st | One dose | 1st | 2nd |
| History of COVID-19 | Probable* | No | Probable | No | No | No | No | No | No | No | No |
| Family history of autoimmune diseases | Sjogren's disease | No | Unknown | Systemic lupus erythematosus | No | No | No | No | No | No | Psoriasis |

\*In-person visit

Those with autonomic symptoms underwent standard autonomic nervous system (ANS) testing including quantitative sudomotor axon reflex testing (QSART) to measure sympathetic post-ganglionic cholinergic sudomotor (sweating) function. Iontophoresis chambers were placed on the forearm, proximal lateral calf, medial ankle, and foot, and sweat volume (µl/mm^2^) was measured on the QSweat Device (WR Medical Electronics, Maplewood, MN). Heart rate variability following 6-8 slow deep breaths for 10 seconds per respiratory cycle was determined as a measure of parasympathetic function. The Valsalva maneuver was used to assess sympathetic and parasympathetic function. Subjects forcibly expired at 40 mm Hg for 15 seconds while heart rate and blood pressure were measured. Tilt table testing was performed with a head-up tilt of 70 degrees for 10 minutes after 20 minutes of supine rest. Changes in heart rate and blood pressure were recorded during head up tilt and after return to supine position. Published normative values were used.^10^ The criteria used to define POTS comprised sustained increase of ≥30 beats per minute from baseline heart rate after 10 minutes in the upright position in the absence of orthostatic hypotension (systolic blood pressure drop of >20 mm Hg, or diastolic blood pressure drop >10 mm Hg). These patients were not on any medications that could affect blood pressure or heart rate, were not in acute pain or deconditioned. They did not have an active infection, diabetes or anemia to explain the orthostasis.

Two skin biopsies using a 3 mm skin punch were removed from the standard locations on the lower leg 10 cm proximal to the lateral malleolus and from the distal lateral thigh, fixed for 16 hours in Zamboni’s fixative, rinsed in 0.1 M Sorensen’s buffer, and cryoprotected for 24 hours. Sections cut 50 μm thick perpendicular to skin surface were immunolabeled for PGP9.5 (Bio-Rad, Hercules, CA) using avidin-biotin complex method and evaluated for SFN as described^11^ and in accordance with established guidelines.^12^

Multiplex fluorescence immunohistochemistry was performed by incubating sections with 5% normal donkey serum (Jackson ImmunoResearch, West Grove, PA) for 1 hour, then overnight at room temperature using 0.5-5 μg/ml mixtures of immunocompatible antibodies (anti-human IgG, anti-human IgM and anti-CD31 (Leica Biosystems; NCL-L-IgG; NCL-L-IgM and PA0414); anti-C1q (Dako; A0136); anti-C4d (Biomedica; BIRC4D and anti-NFH (Aves Labs), followed by a 1 µg/ml mixture of appropriately cross-adsorbed secondary antibodies raised in donkey (Thermo Fisher, Waltham, MA; Jackson ImmunoResearch) and conjugated to one of the following spectrally compatible fluorophores: Alexa Fluor 488, Alexa Fluor 546, and Alexa Fluor 647. Sections were counterstained using 1 µg/ml DAPI (Thermo Fisher) for visualization of cell nuclei and imaged using a Zeiss Imager Z1 microscope (Zeiss, Oberkochen, Germany) equipped with multispectral filters and an Apotome (Zeiss).

## Results

Among twenty-three patients evaluated, ages ranged between 27 to 71 years (median 40 years). Twenty-one (92%) patients were women. None of them had previous neurological illnesses. Five (21%) patients reported skin-flushing, tachycardia and increased blood pressure immediately following the vaccination that lasted less than 30 minutes and resolved completely. All developed neurological symptoms within 21 days of receiving their vaccination, with the median time being four days. Individual clinical characteristics and course of illness can be reviewed in Table 2.

**Table 2:**
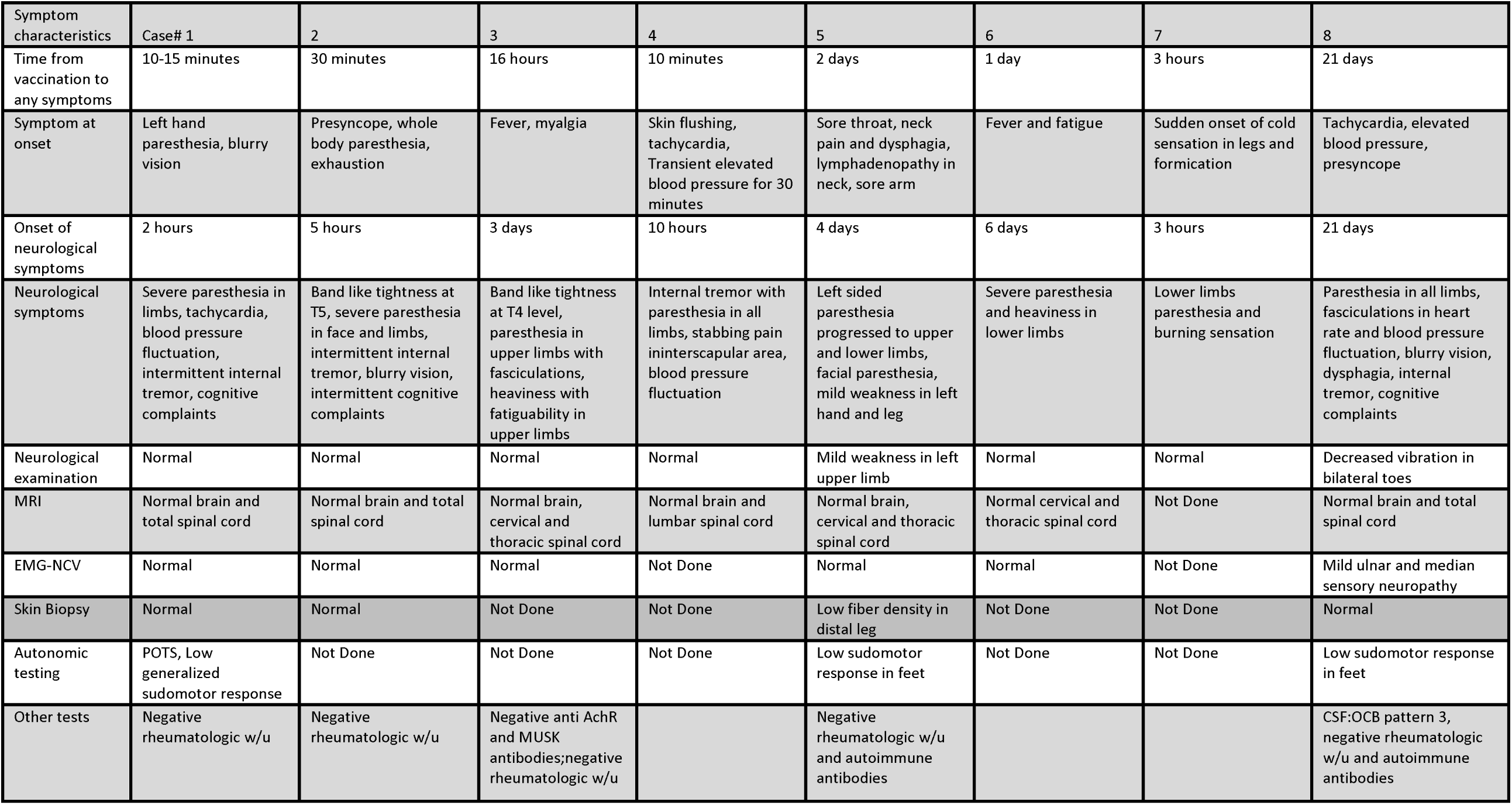
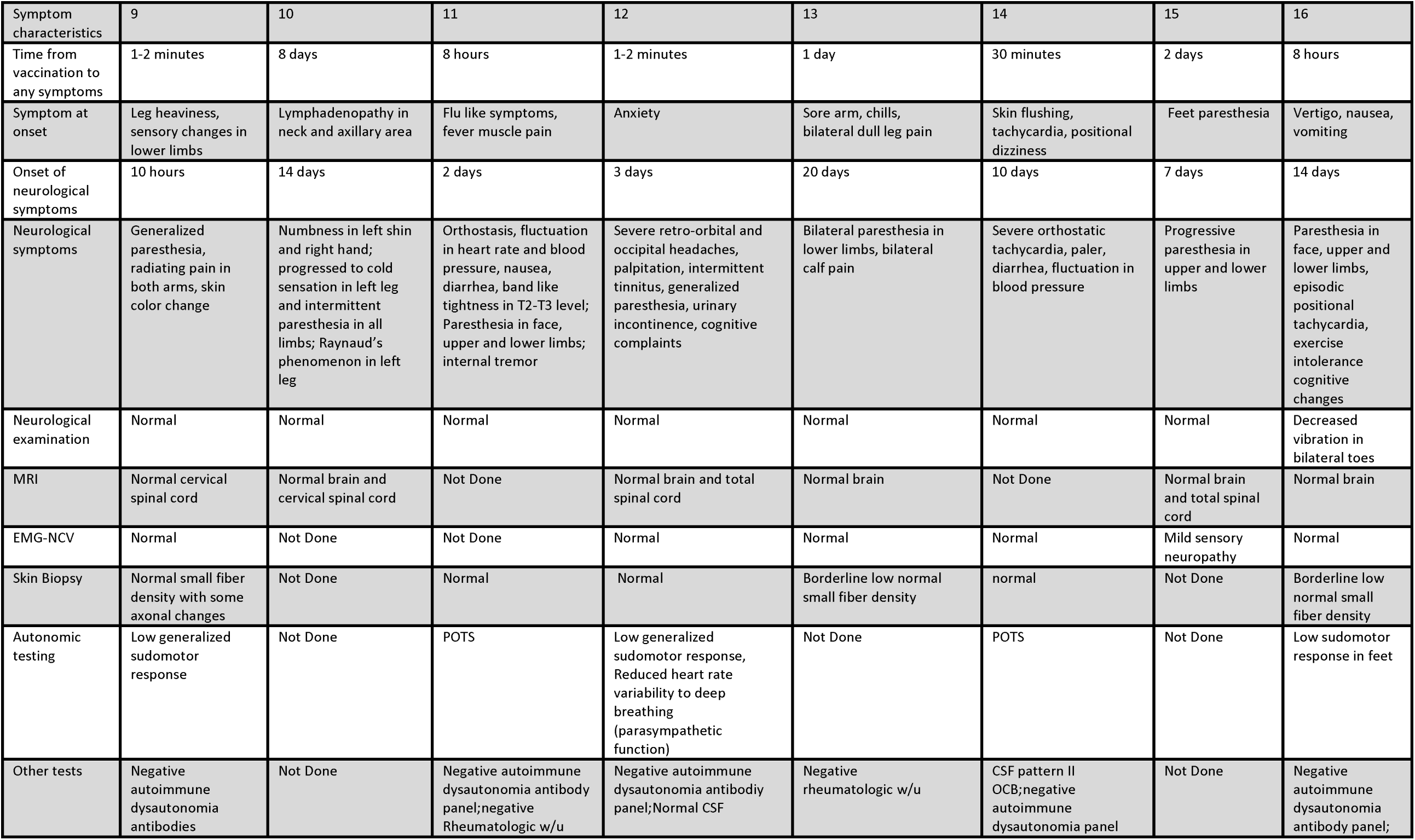
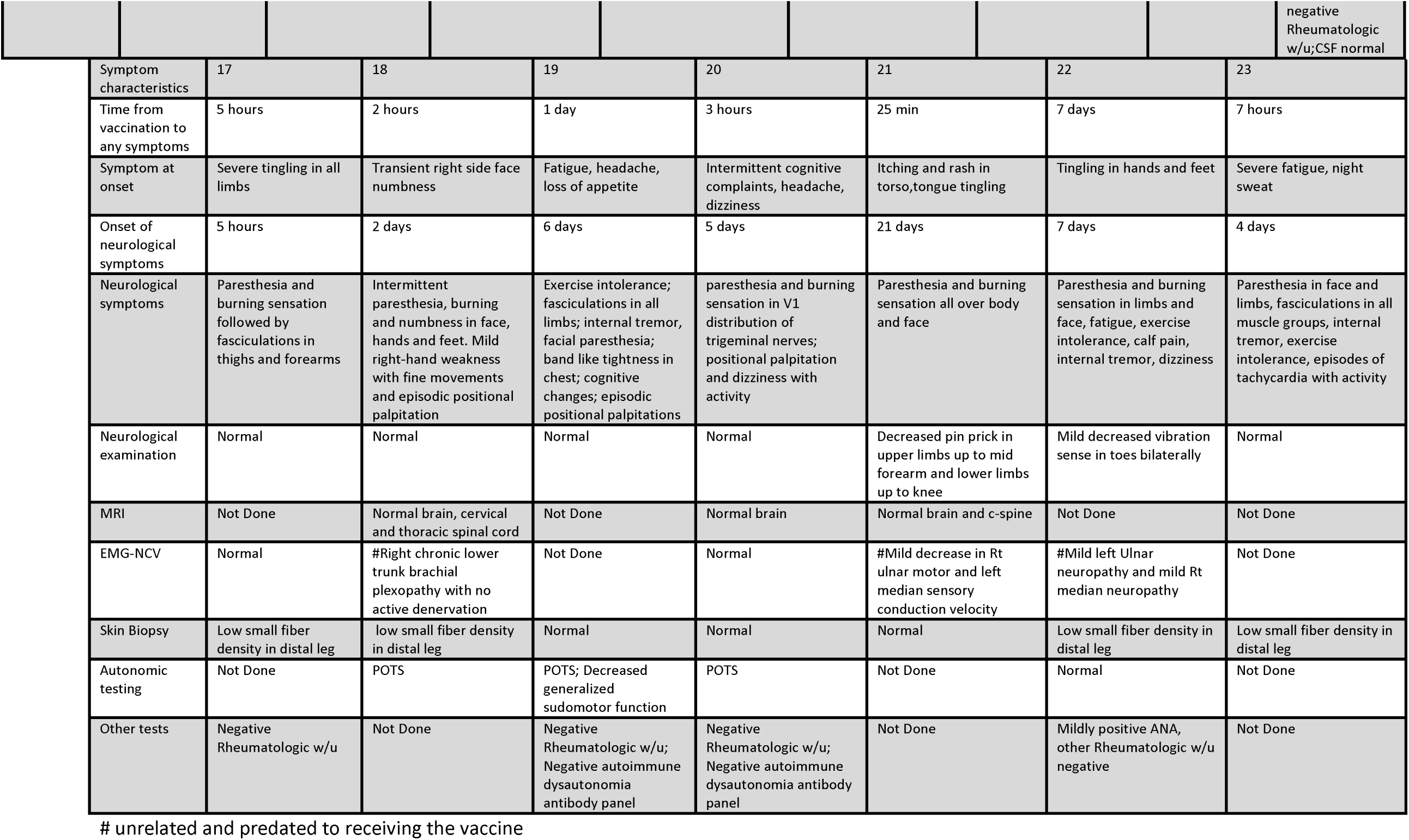
Clinical characteristics and course of illness

The vaccines received included one manufactured by AstraZeneca (ChAdOx1 nCoV19),^13^ one by Janssen (JNJ-78436735), nine by Moderna (mRNA-1273)^14^ and 12 by Pfizer-BioNTech (BNT162b2).^15^ Fourteen participants (65%) developed neurological symptoms following the first dose. Among the 9 (39%) developing symptoms after the 2^nd^ dose, four (17%) reported similar but mild and transient symptoms after their initial vaccination. Two had mild and transient elevation in ALT/AST post-vaccination and two had low-titer ANA and dsDNA antibodies that normalized on subsequent testing. Although none reported symptoms or a confirmed COVID-19 infection before vaccination, we detected antibody to SARS-CoV-2 nucleocapsid protein in one that we attributed to asymptomatic COVID-19.

All participants reported moderate to severe, distal predominant paresthesias and/or burning sensations in both their upper and lower limbs; 9 also had involvement of the face, mouth, and scalp. Neurological examinations were performed in revealed a mild decrease in vibratory sensation in two patients and pinprick pain sensation in another.

Fourteen (60%) patients reported autonomic symptoms, including episodic tachycardia, heat intolerance and new onset Raynaud’s phenomenon. Among the 12 (86%) undergoing ANS testing 11/12 (92%) had abnormal findings. Six (50%) fulfilled the clinical criteria for POTS, seven (58%) had reduced length-dependent sweat production characteristic of SFN. Two (16%) patients had combined POTS and abnormal sudomotor responses. Among 16 patients’ brain and/or spinal cord MRI scans, none identified clinically significant abnormalities.

Among the 16 patients undergoing skin biopsy (Tables 2 and 3), 5 (31%) had subthreshold nerve fiber density fulfilling pathological criteria for SFN. Two (13%) had borderline low density (5.01-10%) at the distal leg site and three (19%) had axonal swellings in the nerve fibers. All had nerve conduction velocities confirming pure small-fiber axonal neuropathy.^16^ Comparing cutaneous deposition of CD31, IgG, IgM, C1q, and C4d between 5 patients and 9 age/sex matched healthy controls revealed more C4d deposition on endothelial cells in all patients (Fig 1). Among the five patients with cerebrospinal fluid (CSF) evaluations, cell counts, protein, glucose and IgG synthesis were normal in all. Two had oligoclonal bands, one with pattern 2 (isolated bands in CSF compared to serum) and another with pattern 3 (both isolated and overlapping bands in CSF compared to serum).

**Table 3a:** Skin biopsy results

|  | 1 | 2 | 5 | 8 | 9 | 11 | 12 | 13 | 14 | 16 | 17 | 18 | 19 | 20 | 21 | 22 | 23 |
| --- | --- | --- | --- | --- | --- | --- | --- | --- | --- | --- | --- | --- | --- | --- | --- | --- | --- |
| Skin fiber density distal leg | 14 | 11.3 | 2.8 | 7.3 | 9 | 13.1 | 8.2 | 6.3 | 17.2 | 8.6 | 4 | 6.2 | 17.1 | 10.5 | 13.1 | 1.2 | 0.5 |
| Skin fiber density distal thigh | Normal | Normal | Normal | Normal | Normal | Normal | Normal | Normal | Normal | Normal | Normal | Not Done | Normal | Normal | Normal | Not Done | Not Done |
| sweat gland nerve fiber density calf | no sweat gland in sample | Not Done | Not Done | 10.6 | Not Done | 16.7 wnl | Not Done | Not Done | 11.1 wnl | 12.6 | Not Done | Not Done | 12..8wnl | 14.9 wnl | 10.7 | Not Done | Not Done |
| sweat gland nerve fiber density thigh | 16.9 wnl | Not Done | Not Done | 18 | Not Done | 18.9 wnl | Not Done | Not Done | 15 wnl | 12.4 | Not Done | Not Done | no sweat gland in sample | no sweat gland in sample | 14.4 | Not Done | Not Done |

**Table 3b:** Small fiber density in skin biopsy

|  | Distal leg 5th percentile |  |
| --- | --- | --- |
| Age | women | men |
| 20-39 | 8 | 5.9 |
| 40-59 | 5.4 | 4.6 |
| 60+ | 3.5 | 3.3 |

**Figure Legend:**
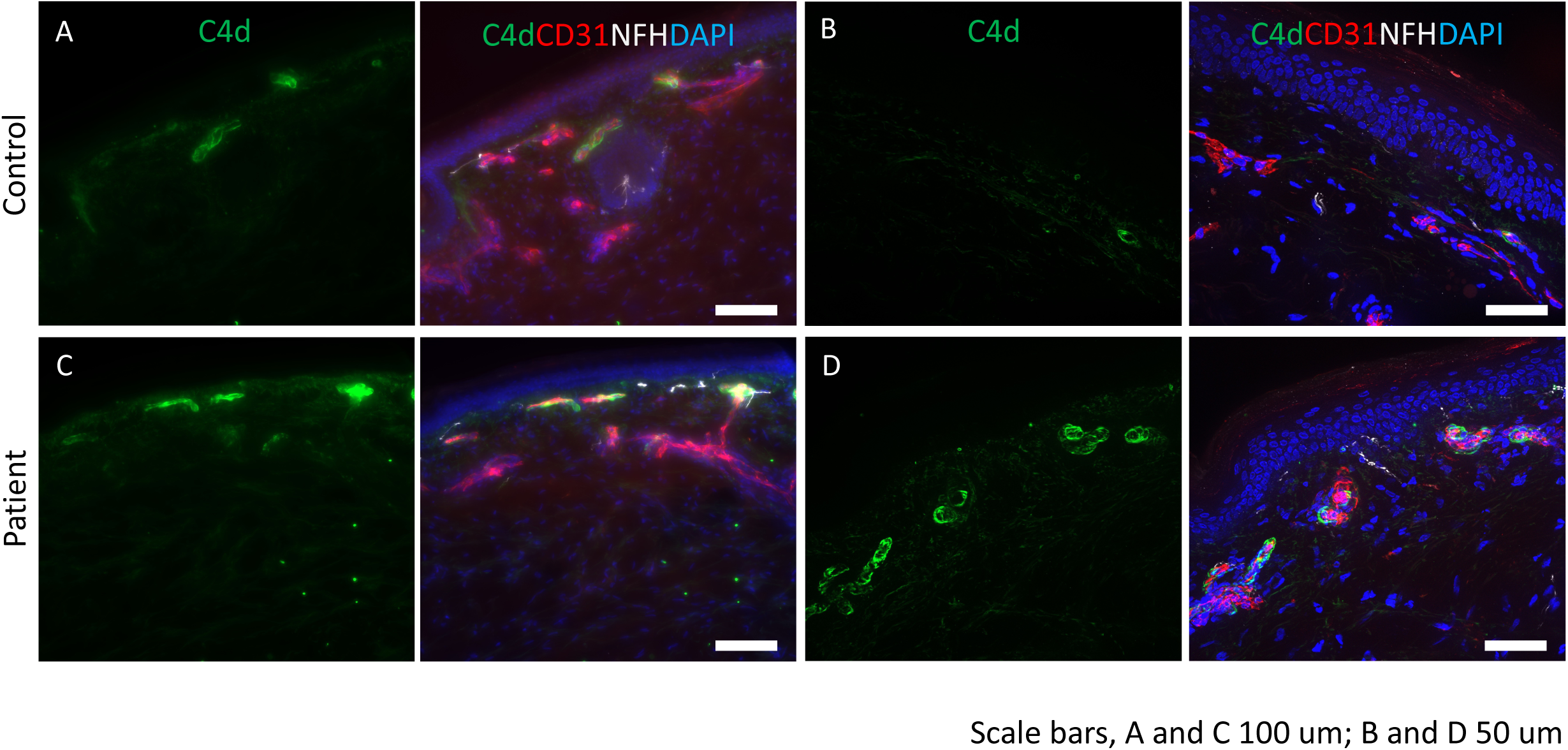
Complement deposition in skin of post-COVID-19 vaccine neuropathy: Immunostaining was performed for C4d (green), endothelial cell marker, CD31 (red) and neurofilament heavy chain, NFH (white). DAPI was used to stain the nuclei. (A and B) control tissues show minimal staining for C4d. CD31 identifies the endothelial cells in the blood vessels. (C and D) deposition of C4d is seen in the endothelial lining of the blood vessels. Scale bars: A and C are 100 mm and B and D are 50 mm.

Treatment with corticosteroid or IVIG had been clinically administered by patients’ treating neurologist or an NIH neurologist consultant. Twelve patients (50%) received oral corticosteroids. Seven of nine (75%) patients who received standard prednisone dosing (0.75-1 mg/kg) for 7 days followed by weekly taper of 20% of the initial dose reported significant symptom improvement after 2 weeks. Of the three treated with a short course of corticosteroid (<7 days) with rapid taper over 1 or 2 weeks, none reported full or expedited improvement after 2 weeks. Three patients who had persistent symptoms of small fiber neuropathy and dysautonomia for 5-9 months were treated with one cycle of IVIg (2g/kg divided over 5 days). Two had been previously treated with corticosteroid with no improvement. In all three, symptoms improved dramatically within 2 weeks of IVIg treatment with complete resolution in one and mild residual symptoms in the other two. Of the 11 patients that never received immunotherapy, seven (64%) had partial recovery, three (27%) have had no improvement, and one (10%) had complete recovery by 12 weeks post-onset as determined by subjective assessment and return to premorbid functional status (Table 4).

**Table 4:** Patient outcome with or without immunotherapy

|  | No recovery<br>after 12 weeks | Partial recovery<br>after 12 weeks | Full recovery after 12<br>weeks | Significant improvement<br>after 2 weeks |
| --- | --- | --- | --- | --- |
| No treatment(n=11) | 3 | 7 | 1 | 0 |
| Corticosteroid long taper(n=9) | 1 | 1 | 7 | 7 |
| Corticosteroid short course(n=3) | 1 | 2 | 0 | 0 |
| IVIg (n=3) | 0 | 0 | 3 | 3 |

## Discussion

This report describes neuropathic symptoms that developed within one month of 1^st^ or 2^nd^ COVID-19 vaccination. All patients in this series reported symptom onset within three weeks of vaccination, 39% developed the symptoms after the second dose, and 17% reported mild/transient symptoms after initial vaccination with full onset after the 2^nd^ dose raise the possibility of immune priming to the spike protein. Although only one had evidence of pre-vaccination COVID-19 infection, cross reactivity between seasonal beta coronaviruses and subunits of the SARS-CoV-2 spike protein might play a priming role.^17^

As an observational study of self-referred patients, it is inherently limited by referral bias. The study is uncontrolled, thus although there was a temporal association of the symptoms to the vaccine, we cannot ascribe it a causative role, although data from the vaccine trials suggests that such manifestations are very rare^4,5^ Case reports link other immune-mediated conditions such as idiopathic thrombocytopenic purpura, acute onset myocarditis and Guillain Barré syndrome to SARS-CoV-2 vaccination, but specificity is also not confirmed.^18^ Not all patients underwent complete evaluations, with some data were collected during clinical care including telemedicine evaluations during the pandemic.

The circumstantial evidence here suggests that in some individuals SARS-CoV-2 vaccination neuropathy may be dysimmune. The fact that participants were screened, and common causes of neuropathy eliminated, the presence of oligoclonal bands in the CSF of two of five participants, deposition of immune complexes on skin biopsy and apparent response to immunotherapy supports possible immune involvement. Plus, almost all patients here were female. Females comprised 69% in one case series of neuropathy incident to COVID-19,^9^ they comprise the large majority of SFN patients,^19^ and of patients with most systemic and organ-specific immune syndromes. Similarly, POTS and multiple types of peripheral neuropathy are well-described after other vaccinations^20^ or illnesses, including brachial plexitis.^21^ The current series extends the hypothesis that COVID-incident neuropathies are dysimmune rather than directly infectious. Postmortem peripheral nerve pathology after fatal COVID-19 infections identified inflammatory perivascular infiltrates of predominantly macrophages without viral antigen in nerves. Detection of interferon stimulated gene MxA in endothelial cells suggested type 1 interferon response.^22^ It is currently unknown if similar mechanisms might contribute to potential vaccine-induced neuropathies.

Various dysimmune large-fiber neuropathies have been temporally associated with SARS-CoV-2 infection including Guillain Barré syndrome, mononeuritis multiplex, plexopathy, multifocal motor neuropathy, and sensory neuropathy, with responsivity to immunotherapies effective for other dysimmune neuropathies reported.^9, 23, 24^ SFN may be associated with long-COVID syndromes.^9, 25, 26^ All our patients had neuropathic symptoms but objective findings of SFN were present in a few patients only. Not all patients with autonomic dysfunction had abnormalities on skin biopsy. In studies of mouse sensory ganglia, small-fiber neurons preferentially display the ACE-2 docking protein for SARS-CoV-2,^27^ consistent with a potential predominance of SFN. Since all SARS-CoV-2 vaccines encode the spike protein^28^, anti-spike protein immune responses may link post-COVID and post-vaccine syndromes. Although the spike protein might interact directly with neurons to mediate the sensory symptoms, anti-idiotypic antibodies to the spike protein immune complex also bind to the ACE-2 receptor.^29^ Dysautonomia during and after SARS-CoV-2 infection is similar but more severe in acutely ill COVID-19 patients^30^ and post-COVID-19 patients.^9, 31^ than in this post-vaccination series.^9, 32^ Alternatively, this response may be non-specific since similar symptoms are reported after other infections and vaccinations. Finally, small fibers lack of myelin which means that they require more energy to maintain axolemmal channels and membrane proteins along the full length of the fiber instead of just at internodes. These fibers also have small amounts of axoplasm and organelles to resupply their distal axons, thus they are preferentially vulnerable to degenerate even from non-specific adversity including diabetes.^5^

Currently, there is insufficient immunological detail. Here, we did not detect the few characterized autoantibodies associated with dysautonomia or neuropathy, but most are not yet characterized. The increased deposition of C4d complement on cutaneous endothelial cells seen here is consistent with immune-mediated activation of the classical complement pathway. C3d and C4d regulatory molecules covalently bind to tissues to formation a cytolytic complex. Endothelial C3d and C4d deposition is reported in SLE and small vessel vasculitis and low blood levels have been reported in some surveys of small-fiber neuropathy.^33, 34^ Cases of seronegative immune-mediated sensory/autonomic neuropathy post-viral infection and post-vaccination implicate memory T-cell responses rather than autoantibodies, as do patients that respond dramatically to high dose corticosteroids but not to plasma-exchange.^7^ And genetic susceptibility to various immune conditions including sensory neuropathy and autonomic dysfunction is documented^37^ Autoantibody generation driven by molecular mimicry and independent immunodysregulation may both contribute.^7^

Enough time has not yet elapsed for the large scale epidemiological studies necessary to confirm or refute causal relations between SARS-CoV-2 vaccination and immune-mediated diseases, thus these results are preliminary. However, although further animal-model and other investigations are needed to confirm the Witebsky postulates,^36^ virtually all preliminary evidence to date supports immune mechanisms. Although some patients may respond to corticosteroids and some severe and refractory cases to IVIg, these agents convey cost and risk of adverse effects and should be used cautiously in well-characterized patients being carefully monitored or be used in the context of a clinical trial. Of note, some patients here improved without immunotherapy. Further studies are needed to determine if there might be a causal relationship between SARS-CoV-2 vaccines and the axonal neuropathies reported here that primarily comprised small fiber polyneuropathy.

## Data Availability

All data produced in the present work are contained in the manuscript

## Acknowledgments

We thank Myoung Hwa Lee, Bryan Smith, Anita Fletcher and Anna Jankowska for valuable help.

